# Usability of three blood-based HIV self-testing devices among men who have sex with men and female sex workers in Yaounde and Douala, Cameroon

**DOI:** 10.1101/2023.10.25.23297570

**Authors:** Awono Noah JP Yves, Justin Ndié, Francis Ateba Ndongo, Onesimus Yongwa, Rogacien Kana, Martial Bonyohe, Plessy Hedgar Mboussam, Tatiana Palisson Avang, Fatima Moulioum, Félicité Tabala Naah, Gutenberg Tchikangni, Audrey Djomo Nzaddi, Alice Ketchaji, Carelle Djofang Yepndo, Gildas Nguemkam, Charles Baudelaire Ndindjock, Brice Seukam, Yagaï Bouba, Ernest Désiré Mvilongo Anaba, Rina Estelle Djoukwe, Serge Billong, Karin Hatzold, Annie Michele Salla, Jérôme Ateudjieu, Anne Cécile Zoung - Kanyi Bissek

## Abstract

**Background:** Blood-based HIV self-testing represents an alternative for increasing screening among key populations or populations with difficult access. In Cameroon, very few studies, on the usability of HIV self-tests based on blood samples, have been carried out among these groups.

**Objective:** The aim of this study was to assess the usability of blood-based HIV self-testing among men who have sex with men (MSM) and female sex workers (FSWs) in the cities of Yaoundé and Douala, Cameroon.

**Materials and Methods:** An observational study was conducted in 17 Community-Based Organisations (CBOs), including 10 MSM and 07 FSWs in Yaoundé and Douala from 11 to 22 June 2022. The study population consisted of HCV and MSM aged 21 years and over who agreed to participate in the study. After they were recruited consecutively in their respective CBOs, they received counselling, unassisted HIV blood self-testing and condoms. Data was collected using an administered questionnaire. Three HIV blood self-testing devices were used in the study: Mylan HIV Self-Test, Sure Check HIV Test® (Chembio Diagnostics Inc), Check Now HIV Self-Test(Abbott™ Point Of Care). Analysis was conducted using SPSS 23 software with a 95% confidence level.

**Results:** Of 817 participants who completed the HIV blood self-test, just over half were TS 459(56.2%); the median age was 27 years (IQR: 22 years - 34 years) and the 25-49 age group was most represented 482(59.0%). One participant in ten (10%) had never been tested for HIV. However, 98.6% of participants agreed to use the HIV blood self-test and the vast majority (97.1%) followed the steps for carrying out the HIV blood self-test. An MSM was 4 times more likely to pass an HIV blood self-test than a TS (aOR= 4.01; 95% CI: 1.181-13.625; p=0.026). Similarly, TS and MSM who used the Abbott Check Now HIV Self-Test (aOR= 3.85; 95% CI: 1.246-11.908; p=0.019) and Chembio Sure Check HIV Test (aOR= 2.83; 95% CI: 1.072-7.720; p=0.036) were respectively 3.8 and 2.8 times and more likely to pass their self-test than those who used the Mylan blood self-test. Agreement between a participant’s HIV blood self-test result and Abbott-trained investigator-observers was moderate (κ=0.485; CI95% (0.359-0.610); p=0.001) while agreement with Chembio and Mylan was respectively low (κ=0.329; CI95% (0.203-0.454); p=0.001) and very low (κ=0.194; CI95%(0.05-0.329); p=0.001).

**Conclusion:** HIV blood self-testing is acceptable and usable by key populations in Cameroon. Although usability was limited by problems in interpreting results and incorrect disposal of waste t by key populations, a blood-based HIV Self-Test, with moderate concordance, proved suitable for unassisted use in key populations, what could help improve HIV prevention interventions.

## Introduction

In every region of the world, key populations -men who have sex with men (MSM), female sex workers (FSWs), drug users and transgender people- and their partners are much more affected by HIV infection than the general population. In sub-Saharan Africa specifically, key populations and their partners accounted for 51% of new HIV infections in 2021. Given the importance of MSM and MSM-attached populations, where routine HIV screening strategies have produced mixed results, the WHO recommends implementing new strategies such as HIV self-testing (HIVST) [1]. Hence the prequalification of the first salivary HIV self-testing kit by the WHO in 2017. The first HIV blood self-test was prequalified in 2019, followed by 4 other blood self-tests between 2019 and 2022. In terms of institutional support, formal HIV self-testing regulations have existed in many African countries since 2017, whereas they are still being developed or even informal in other countries [2]. However, in sub-Saharan Africa, apart from the Democratic Republic of the Congo (DRC), the Central African Republic (CAR) and Benin, very few French-speaking countries have shown concern for blood-based HIV self-testing. In the DRC, evidence has been produced in favour of the feasibility and performance of blood self-testing in the general [3] and key populations such as FSWs [4] or vulnerable populations such as adolescents [5]. In general, the majority of users were able to understand the instructions for use, identify the different components of the self-test kit, perform the test until a valid test was obtained, and good interpretation as well. Many of the advantages highlighted by these studies, such as confidentiality, speed, and efficiency, were pointed out by users, despite some difficulties encountered in using the lancet and misinterpreted results. These studies suggest that blood self-testing could be considered as an additional self-testing option for key populations. In Cameroon, in 2018, HIV prevalence in the general population (15-49 years) was estimated at a rate of 2.7% [6]. The percentage among MSM and FSWs was 20.6% and 24.3% respectively [7]. In addition to the burden of HIV, these populations are afraid of stigmatisation and discrimination as a result of taking part in conventional HIV screening programmes. A study conducted in Douala-Cameroon, has shown that only 48% of MSM had already been exposed to HIV prevention interventions [8]. Some research carried on the acceptability of salivary HIV self-tests among MSM and FSWs in Yaoundé, has shown a high level of acceptability and has led to the adoption of salivary HIV self-testing among MSM and FSWs in Cameroon [9]. If blood-based HIV self-tests are to be introduced in this country as a complement to the already-approved saliva self-test, there must first of all be evidence of their usability. To date, no locally conducted and published study has shed any light on this strategic option. To sum up, the aim of this study was to assess the usability of HIV self-testing among men who have sex with men and female sex workers in Yaoundé and Douala. The study included compliance with the steps involved in carrying out the HIV blood self-test and interpreting the results.

## Methodology

### Ethical considerations

This protocol obtained a favourable opinion from the National Ethics Committee for Human Health Research of Cameroon (reference numbers 2022/04/1448/CE/CNERSH/SP) and from the Operational Health Research Division of the Ministry of Health of Cameroon (Administrative Research Authorisation reference numbers 031-16-22). A written informed consent was obtained from all participants. All data collected was treated confidentially and stored within the limits permitted by law.

### Type of study

A cross-sectional and observational study was conducted from 11 to 22 June 2022 in 17 community-based organisations (CBOs) in Yaoundé and Douala.

### Selection of the study population

The study population consisted of MSM and FSWs, aged 21 years and over, declaring unknown HIV status or HIV-negative for more than three months, fluent in French and/or English and usually receiving HIV testing services offered by CBOs involved in the implementation of HIV control activities. Any participants who were on PrEP (pre-exposure prophylaxis) or antiretroviral treatment, had a known HIV-positive status and/or had extenuating conditions (e.g., acute illness) that might interfere with the study process were automatically excluded.

### Site selection

The study was carried out in Cameroon, a Central African country with a population of around 25 million. The study was conducted in the two major Cameroonian cities namely Yaoundé (the capital city) and Douala (the economic capital). They were chosen because of the size of their Community-Based Organisations (CBOs), key populations and their high “hot spot” densities. In each study city, the CBOs were selected based on the size of the target effectively covered and the acceptance of the study by their leaders.

### Sample size and sampling

The sample size was calculated according to the criteria recommended by the WHO [10] taking into account the proportion of HIV testing observed in each target group in previous national or regional studies. The minimum sample size adjusted for the 15% non-response rate was estimated at 785 study participants (423 TS and 362 MSM), including 262 for each HIV self-testing device.

### Recruitment of MSM and sex workers

MSM were recruited by leader mothers, innkeepers, and gatekeepers in the community and in the hot spots. They were recruited consecutively until the planned sample size for each type of HIV blood self-test was reached. MSM were mobilised by peer educators and community leader fathers. They were also recruited consecutively until the planned sample size for each type of HIV blood self-test was reached.

### Data collection

Data collection was carried out by a research team consisting of twenty healthcare staff who had been trained to perform blood self-tests (laboratory technicians and nurses). The study ran for 9 months from November 2021 to July 2022 and included eight (08) days of data collection from 11 to 22 June 2022. In each CBO selected for the survey, a setting was set up to conduct the data collection and HIV blood self-test procedures in complete confidentiality. To begin with, data collectors introduced themselves to every participant. After that authorisations for the study were distributed and its purpose highlighted. Then, participants were given an information leaflet and their informed consent was obtained. Information about their eligibility for the study was also collected. Next, the blood self-test kit was given to the recruited participant to carry out the blood self-test procedure under observation. The tests were done on site and the results were reported by participants and two trained investigator-observers in a row. CBO managers and staff provided pre- and post-test counselling to each participant. Once the self-testing process was complete, every investigator-observer carried on with the administration of the questionnaire to one participant at atime. As there were no available standardised questionnaires about the usability of HIV blood self-tests at the time of the study, a product-specific semi-structured questionnaire was developed (Text S1) based on WHO documentation on prequalification [11-13]. The questionnaire was tested with a sample of 43 participants (19 MSM and 24 FSWs) for each HIV self-test that had to be used. A semi-structured questionnaire, set up on the tablets via the Kobo Collect application, was administered by observers. The questionnaire included an observation checklist of the HIV self-testing process, test results, waste management and participants’ experiences with the device and instructions for use. The observer systematically provided post-test counselling and condoms to study participants after they had completed the test and read the results. MSM/FSWs with a reactive test were referred to their CBO’s supervising health facility for confirmation of their test result in accordance with the national algorithm for HIV testing and management where appropriate.

### Type of blood-based Self-test devices

Three blood-based HIV Self-Testing (HIVST) devices were used in the study: Mylan® HIV Self-Test (Atomo Diagnostics Pvty. Ltd), Sure Check® HIV Self-Test (Chembio Diagnostic Systems, Inc), Check Now™ HIV Self-Test (Abbott Rapid Diagnostics Jena GmbH). Each HIV Self-Test device included the manufacturer’s instructions for use and other kit components. No additional user aids, demonstrations or assistance were provided to participants.

## Data processing and statistical analysis

After data was collected with the KoboCollect application deployed through the KoboToolbox platform, it was imported into SPSS (version 23) for analysis using descriptive statistics and multivariate regression models. For this study, usability of the blood self-test was defined as the proportion of MSM/STDs who were able to perform self-testing steps, interpret the result and dispose of the waste in exact compliance with the manufacturer’s instructions. Variables that were statistically significant (p-value <0.05) in the univariate analysis were included in the multivariate logistic regression. Multivariate logistic regression analysis was used to identify factors associated with successful self-testing. The results of the analysis are presented as adjusted odds ratios (aOR) with 95% confidence intervals (CI) and interpreted as the odds of successful completion of the self-test among non-professional users who have been exposed or not to the associated factor. Variables included in the multivariable logistic regression analysis were city, type of key population, age, sex, education, employment, living status, ever HIV tested, and type of blood based HIVST kit. Self-test success or self-test usability was calculated as the percentage of participants who correctly performed the key steps and obtained an interpretable result. Tests that did not produce a control line were identified as “Invalid” and reported as a failure.

Agreement between reactive (positive) and non-reactive (negative) HIV self-test results was measured using Cohen’s Kappa based on agreement rates between reactive and non-reactive self-test results interpreted by study participants and observing healthcare staff. This analysis excluded incomplete self-tests, invalid results interpreted by healthcare staff and inconclusive (uncertain) results interpreted by study participants.

## Results

### Demographic characteristics and HIV testing history of MSM and FSW

Of the 817 participants who agreed to be tested for HIV, 56.3% were living in the city of Douala; almost all of them were Cameroonian (98.2%); 56.2% were MSM; the median age was 27 years and the 25-49 age group was the most represented (59%); the majority of participants had completed secondary education (58.8%); the unemployed accounted for just under half of the sample (43.3%); the vast majority of participants were single (77.8%); one in ten participants (10%) had never been tested for HIV before the test was carried out as part of this study. A wide disparity between the two sub-populations was observed (17.9% of MSM compared with 3.9% of MSM). 280 Chembio Sure Check HIV Test®, 272 Abbott™Check Now HIV Self-Test and 265 Mylan® HIV Self-Test were used for this study (Table 1).

**Table I:**
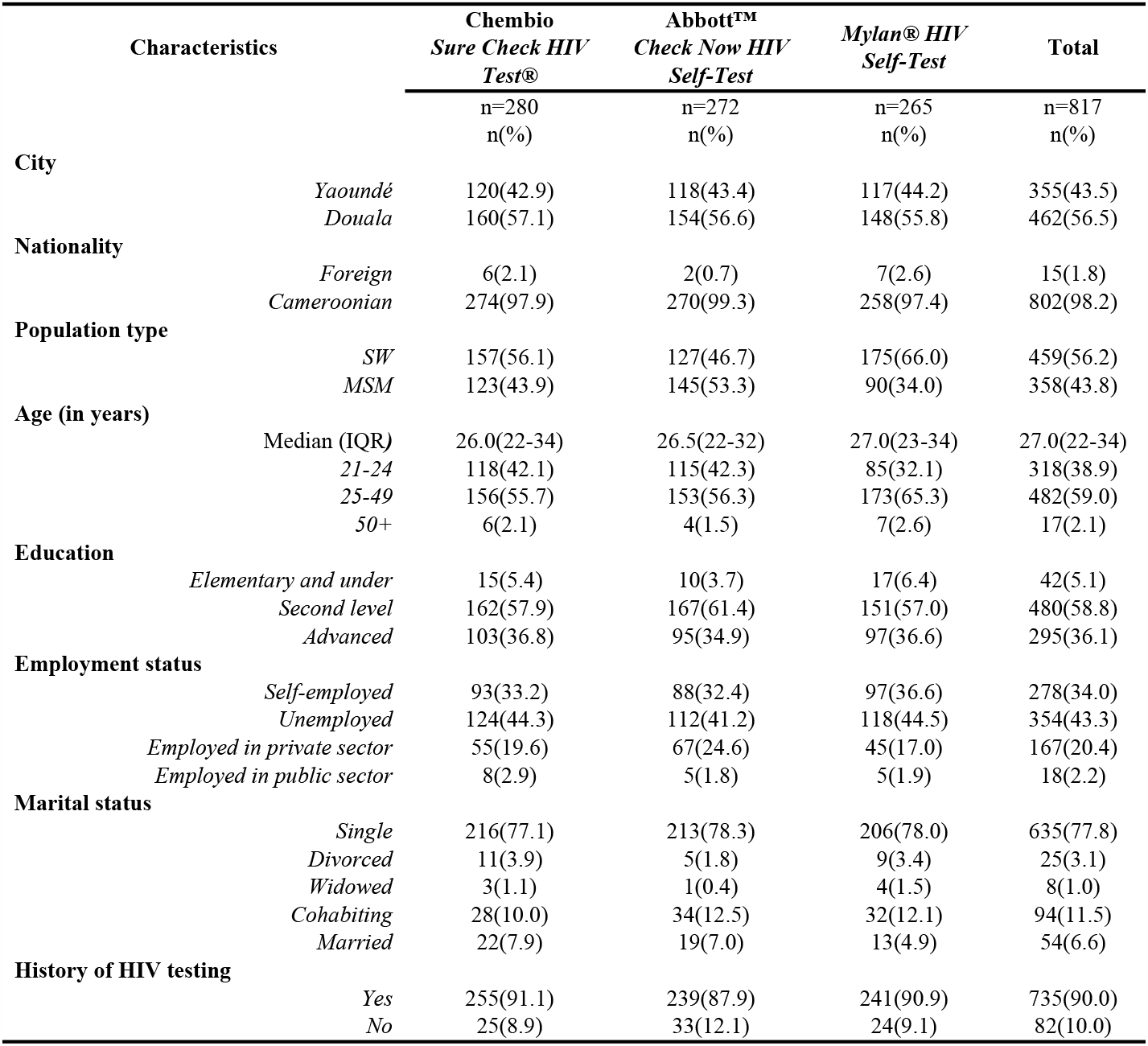
Distribution of TS and MSM according to sociodemographic characteristics and HIV testing history.

| Characteristics |  | Chembio<br>*Sure Check HIV*<br>*Test®* | Abbott™<br>*Check Now HIV*<br>*Self-Test* | *Mylan® HIV*<br>*Self-Test* | Total |
| --- | --- | --- | --- | --- | --- |
|  |  | n=280<br>n(%) | n=272<br>n(%) | n=265<br>n(%) | n=817<br>n(%) |
| City | *Yaoundé* | 120(42.9) | 118(43.4) | 117(44.2) | 355(43.5) |
|  | *Douala* | 160(57.1) | 154(56.6) | 148(55.8) | 462(56.5) |
| Nationality | *Foreign* | 6(2.1) | 2(0.7) | 7(2.6) | 15(1.8) |
|  | *Cameroonian* | 274(97.9) | 270(99.3) | 258(97.4) | 802(98.2) |
| Population type | *SW* | 157(56.1) | 127(46.7) | 175(66.0) | 459(56.2) |
|  | *MSM* | 123(43.9) | 145(53.3) | 90(34.0) | 358(43.8) |
| Age (in years) | Median (IQR) | 26.0(22-34) | 26.5(22-32) | 27.0(23-34) | 27.0(22-34) |
|  | *21-24* | 118(42.1) | 115(42.3) | 85(32.1) | 318(38.9) |
|  | *25-49* | 156(55.7) | 153(56.3) | 173(65.3) | 482(59.0) |
|  | *50+* | 6(2.1) | 4(1.5) | 7(2.6) | 17(2.1) |
| Education | *Elementary and under* | 15(5.4) | 10(3.7) | 17(6.4) | 42(5.1) |
|  | *Second level* | 162(57.9) | 167(61.4) | 151(57.0) | 480(58.8) |
|  | *Advanced* | 103(36.8) | 95(34.9) | 97(36.6) | 295(36.1) |
| Employment status | *Self-employed* | 93(33.2) | 88(32.4) | 97(36.6) | 278(34.0) |
|  | *Unemployed* | 124(44.3) | 112(41.2) | 118(44.5) | 354(43.3) |
|  | *Employed in private sector* | 55(19.6) | 67(24.6) | 45(17.0) | 167(20.4) |
|  | *Employed in public sector* | 8(2.9) | 5(1.8) | 5(1.9) | 18(2.2) |
| Marital status | *Single* | 216(77.1) | 213(78.3) | 206(78.0) | 635(77.8) |
|  | *Divorced* | 11(3.9) | 5(1.8) | 9(3.4) | 25(3.1) |
|  | *Widowed* | 3(1.1) | 1(0.4) | 4(1.5) | 8(1.0) |
|  | *Cohabiting* | 28(10.0) | 34(12.5) | 32(12.1) | 94(11.5) |
|  | *Married* | 22(7.9) | 19(7.0) | 13(4.9) | 54(6.6) |
| History of HIV testing | *Yes* | 255(91.1) | 239(87.9) | 241(90.9) | 735(90.0) |
|  | *No* | 25(8.9) | 33(12.1) | 24(9.1) | 82(10.0) |

### Usability of the HIV blood self-test by study participants

The vast majority of participants (97.1%) followed the steps for performing the self-test; the errors commonly made while using the self-test were, in order of occurrence: incorrect volume of blood drawn (87.9%); insufficient collection/addition of blood to the well (29.9%), particularly with Chembio (39.3%) and Mylan (37.4%); failure to dispose of waste according to the leaflet (23.4%), particularly with Chembio (24.3%) and Mylan (27.5%); incorrect positioning of the self-test device (20.4%), particularly with Chembio (22.9%) and Mylan (28.7%) (Table 2).

**Table II:** Ability to perform HIV self-testing in exact compliance with the leaflet.

| Characteristics | Chembio®<br>*Sure Check HIV*<br>*Self-Test* | Abbott™<br>*Check Now HIV*<br>*Self-Test* | *Mylan® HIV*<br>*Self-Test* | Total |
| --- | --- | --- | --- | --- |
|  | n=279<br>n(%) | n=272<br>n(%) | n=263<br>n(%) | n=814<br>n(%) |
| **Completeness of self-test steps** |  |  |  |  |
| *Success* | 273(97.8) | 268(98.5) | 249(94.7) | 790(97.1) |
| *Failure* | 6(2.2) | 4(1.5) | 14(5.3) | 24(2.9) |
| **Frequent errors made during self-testing process** |  |  |  |  |
| *Incorrect volume of blood drawn (tube over/underfilled)* | 246(91.1) | 226(86.9) | 217(85.4) | 689(87.9) |
| *Insufficient blood collected/added to the well* | 110(39.3) | 32(11.8) | 99(37.4) | 241(29.5) |
| *Waste not disposed of in accordance with instructions* | 68(24.3) | 50(18.4) | 73(27.5) | 191(23.4) |
| *Incorrect positioning of the device* | 64(22.9) | 27(9.9) | 76(28.7) | 167(20.4) |

Regarding agreement in the interpretation of results between participants and the healthcare workers trained as HIV testing counsellors, overall, the percentage of positive agreement was 90.9% and the percentage of negative agreement was 81.7%. The total percentage of agreement was 82.3%. However, when controlling the random effect, this total percentage of agreement dropped to 35.1%, indicating low overall agreement (κ=0.351; 95% CI (0.274-0.427); P=0.001). Notwithstanding, disparities were identified depending on the blood self-test device used. Indeed, agreement between a participant’s (MSM or TS) HIV blood self-test result and investigator-observers trained with **Abbott™ Check Now HIV Self-Test** was moderate (κ=0.485; 95% CI (0.359-0.610); P=0.001) while agreement with **Chembio Sure Check HIV Test®** and **Mylan® HIV Self-Test** was respectively low (κ=0.329; 95% CI (0.203-0.454); P=0.001) and very low (κ=0.194; 95% CI (0.05-0.329); P=0.001) with percentages of total agreement corrected by the random effect of 32.9% and 19.4% (Table 3).

**Table III:**
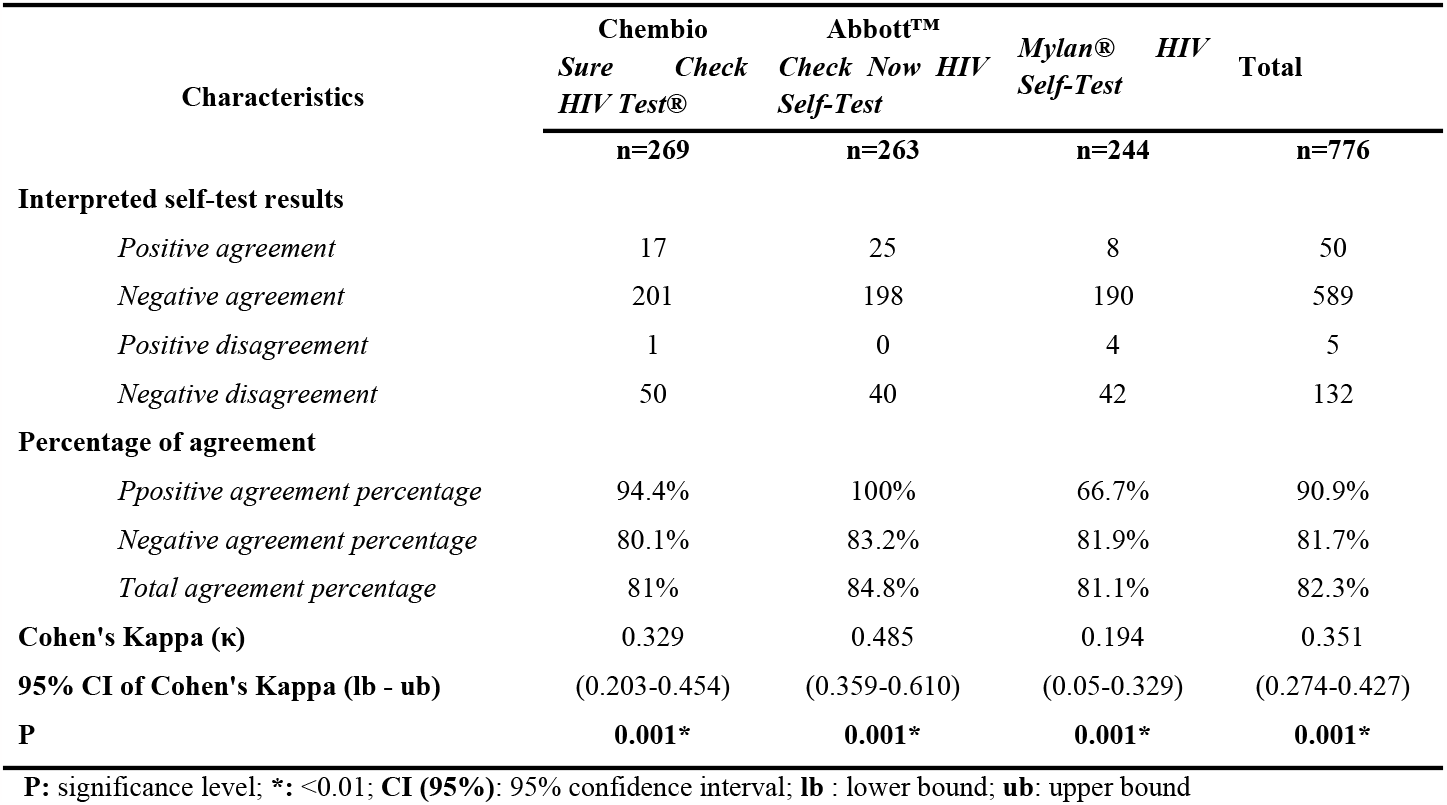
Agreement between HIV blood self-test results interpreted respectively by participants trained health workers.

### Factors associated with successful HIV blood self-testing

Multivariate logistic regression analysis (Table 4) showed a significant positive association between successful self-testing with Abbott Check Now HIV Self-Test (aOR= 3.85; 95% CI: 1.246-11.908; p=0.019), Chembio Sure Check HIV Test (aOR= 2.83; 95% CI: 1.072-7.720; p=0.036) and being MSM (aOR= 4.01; 95% CI: 1.181-13.625; p=0.026). Multivariable analysis also confirmed a significant negative association between self-test success and the city of residence (aOR = 0.27; 95% CI: 0.110-0.676). Age over 30 years, which was nearly negatively associated with successful self-testing in the univariate analysis, was not confirmed in the multivariate analysis after adjustment with other variables. In fact, MSM were 4 times more likely to succeed in self-testing than an FSW. Similarly, FSW and MSM who used the Abbott and Chembio blood self-test were respectively 3.8 and 2.8 times more likely to pass their test than those who used the Mylan blood self-test. In addition, the probability of carrying out their blood self-test successfully was 73% lower for participants living in the city of Yaoundé than for those living in the city of Douala.

**Table IV:**
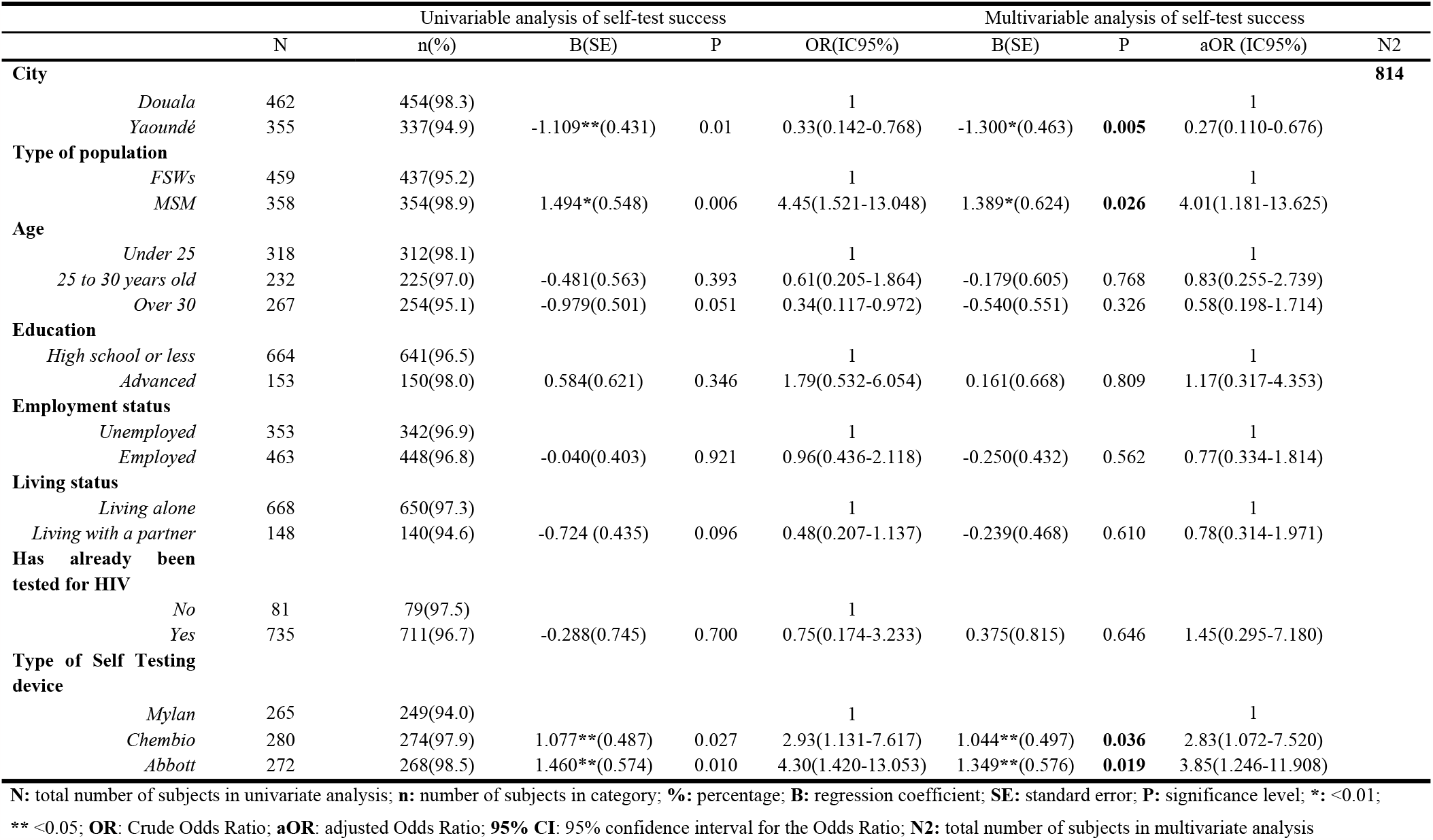
Factors associated with successful completion of all stages of blood self-testing.

## Discussion

The HIV epidemic in Cameroon is widespread among key populations such as sex workers and men who have sex with men, with national prevalence rates of 5.6% and 10.4% respectively. The Cameroon government is committed to achieving the UNAIDS 95-95-95 targets and to putting an end to AIDS by 2030. Innovations in HIV testing, namely testing by a non-professional provider and self-testing, have been implemented in Cameroon since 2015 (when community-based testing was validated by CBOs) and 2022, respectively, with the aim of increasing access to routine services for people who have not been reached yet. Available data in Cameroon shows that these screening innovations are effective in reaching undiagnosed people, those getting screened for the first time or those who would not like to get screened or come to health facilities as well (Spectrum programmatic data). The aim of this study was to assess whether blood-based HIV testing kits could be used appropriately by unassisted non-professionals in a context of low HIV prevalence in Cameroon. The ease of use of blood-based HIV tests by non-professional users raised concerns about the potentially infectious nature of waste from the procedure compared with oral fluid-based HIV self-tests. The results of this study confirmed that unassisted blood-based HIV self-testing was perfectly feasible and that the success of self-testing was significantly associated with the fact of being MSM and with brand too. Indeed, the Abbott self-test had a 223% and MSM a 491% greater chance of passing the steps involved in carrying out the test, compared with Mylan and FSWs respectively. In the same vein, these results are similar to those of previous studies on HIV blood screening kits conducted in the Democratic Republic of the Congo, Thailand, Central African Republic and South Africa and Canada [3, 5, 14-16]. The study showed no association with employment status unlike Bao, Majam, Green, Tran, Hung, Anh Luong Que et al.[17] who found an inverse association between the type of self-test and self-test success. The proportion of MSM who had never been tested for HIV prior to this study was 17.9%, which is more than three times higher than that found among MSM in Nairobi [18] but half lower than that found in Canada among student population [19]. Despite the feasibility of blood-based HIV self-tests, the study revealed errors in the process of obtaining the blood sample or transferring it to the test well, failure to dispose of waste according to the instructions, and incorrect positioning of the device; these errors suggest the need to modify current rules so as to make the use of self-tests easier and easier These findings confirm those of previous studies in which most errors occurred during the sample collection stage by users of finger-prick HIV tests [15-17, 19]. It is worth mentioning that Abbott recorded the fewest errors. It was also found that the correct interpretation of HIV self-test blood test results by non-professional users was, based on the 95% confidence intervals of Cohen’s Kappa statistic, very low to low, low to moderate and moderate to high for Mylan, Chembio and Abbott respectively, compared with interpretation by trained healthcare staff. This suggests that Abbott is probably appropriate for use in key populations on an unassisted basis. Difficulties in interpreting results have also been reported in the literature, with differences between the different approaches [3-5, 14,20]. A systematic review reported that in studies using the unassisted approach, reactive results were often misinterpreted as non-reactive [20]. Misinterpretation of positive results could have a negative impact on the control of the HIV epidemic [21], as HIV self-testing is considered a triage test [1, 22].

## Conclusion

This study showed a high level of usability (98%) of HIV blood self-tests among key populations (MSM and FSWs) in the cities of Yaoundé and Douala. Despite the fact that usability was restricted by results related issues and inappropriacy of waste disposal by key populations, the Abbott blood self-test, with a moderate agreement, proved suitable for unassisted use in key populations. These results are essential as they could help promote the spread of blood-based HIV testing in addition to oral fluid-based HIV testing so as to offer a greater choice of products to potential testees, thereby increasing uptake of HIV testing and access to essential HIV prevention such as pre-exposure prophylaxis (PrEP) and treatment services. The COVID-19 pandemic has made access to a range of affordable blood- and oral fluid-based HIV tests more crucial than ever. Given the fact that restrictions are gradually lifted and people are more and more getting access to safe and personal care, HIV self-tests are an essential tool for accelerating catch-up HIV testing among the plethora of people who have not benefited from it yet.

## Data Availability

The datasets used and/or analyzed during the current study are available from the corresponding author on reasonable request.

## Acknowledgments

We are grateful to all the study participants for their contributions. We would also like to express our gratitude to Mohammed Majam, for his collaboration and commitment in implementing the study. We also thank Cameroon’s Ministry of Public Health (MoPH), Population Service International (PSI) - STAR Project and Cameroon Social Marketing Association, Cameroon (ACMS) for providing the necessary and accurate resources.

## References

[1] OMS. Lignes directrices sur l’autodépistage du VIH et la notification aux partenaires : supplément aux lignes directrices unifiées sur les services de dépistage du VIH [Guidelines on HIV selftesting and partner notification: supplement to consolidated guidelines on HIV testing services]. 2018. Genève. https://creativecommons.org/licenses/by-nc-sa/3.0/igo/

[2] World Health Organization & UNITAID. Technology Landscape, HIV rapid diagnostic tests for self-testing, December 2016, Semi-annual update. Geneva : WHO & UNITAID, 2016. http://www.who.int/hiv/pub/vct/hiv-self-testing-2016-december-edition/en/ (last access March 2017).

[3] Grésenguet G, Longo JD, Tonen-Wolyec S, Mboumba Bouassa RS, Belec L. Acceptability and Usability Evaluation of Finger-Stick Whole Blood HIV Self-Test as An HIV Screening Tool Adapted to The General Public in The Central African Republic. Open AIDS J. 2017; 11:101–118

[4] Tonen-Wolyec S, Batina-Agasa S, Longo JDD, Mboumba Bouassa RS, Bélec L. Insucient education is a key factor of incorrect interpretation of HIV self-test results by female sex workers in Democratic Republic of the Congo: a multicenter cross-sectional study. Medicine. 2019 Feb;98(6):e14218.

[5] Tonen-Wolyec S, Batina-Agasa S, Muwonga J, Mboumba Bouassa R-S, Kayembe Tshilumba C, Bélec L. Acceptability, feasibility, and individual preferences of blood based HIV self-testing in a population-based sample of adolescents in Kisangani, Democratic Republic of the Congo. PLoS One. e0218795;14(7).

[6] Institut National de la Statistique et ICF International. Rapport de l’Enquête Démographique et de Santé et à indicateurs multiples du Cameroun 2016. 2018. Calverton, Maryland, USA : INS et ICF International https://dhsprogram.com/pubs/pdf/fr260/fr260.pdf

[7] Comité National de lutte contre le SIDA. (2017); 2016 Integrated Biological and Behavioral Survey (IBBS) Report among Key Populations in Cameroon : Female sex workers and men who have sex with men

[8] Holland CE, Papworth E, Billong SC, Kassegne S, Petitbon F, Mondoleba V, et al. (2015) Access to HIV Services at Non-Governmental and Community-Based Organizations among Men Who Have Sex with Men (MSM) in Cameroon: An Integrated Biological and Behavioral Surveillance Analysis. PLoS ONE 10(4): e0122881. 10.1371/journal.pone.0122881

[9] Billong SC, Eyebe S, Mossus T, Njindam Mfochive I, Tamoufe U, Fako G, Baral S, Zoung-Kanyi AC, Kamgno J. Feasibility of demedicalization of HIV diagnosis in Cameroon: experimentation with HIV self-test in “key populations”.Health Sci. Dis: Vol 22 (2) February 2021 pp 23–27

[10] Stanley Lemeshow S, Hosmer DWJr, Klar J and Lwanga SJ. Adequacy of Sample Size in Health Studies. WHO, published by John Wiley & sons, 1991

[11] WHO Prequalification of In Vitro Diagnostics PUBLIC REPORT Product: CheckNOW HIV SELF TEST WHO reference number: PQDx 0481-032-00. 2022. available on https://extranet.who.int/pqweb/WHOPR/public-report-checknow-hiv-self-tespqdx-0481-032-00

[12] WHO Prequalification of In Vitro Diagnostics PUBLIC REPORT Product: SURE CHECK HIV SELF TEST WHO reference number: PQDx 0054-006-01. 2019. available on https://extranet.who.int/pqweb/content/public-report-sure-check-hiv-self-testpqdx-0054-006-01

[13] WHO Prequalification of In Vitro Diagnostics PUBLIC REPORT Product: MYLAN HIV SELF TEST WHO reference number: PQDx 0320-090-00. 2019. available on https://extranet.who.int/pqweb/content/public-report-mylan-hiv-self-testpqdx-0320-090-00

[14] Tonen-Wolyec S, Tshilumba CK, Batina-Agasa S, Djang’eing’a RM, Hayette MP, Bélec L. Comparison of practicability and effectiveness between unassisted HIV self-testing and directly assisted HIV self-testing in the Democratic Republic of the Congo: a randomized feasibility trial. BMC Infect Dis. 2020 Nov 11;20(1):830. pmid:3317670

[15] Smith P, Wallace M, Bekker LG. Adolescents’ experience of a rapid HIV self-testing device in youth-friendly clinic settings in Cape Town South Africa: a cross-sectional community based usability study. Journal of the International AIDS Society 2016, 19:21111| 10.7448/IAS.19.1.21111

[16] Galli RA, Tian JMLH, Sumner-Williams M, McBain K, Stanizai E, Tharao W, et al. An observed, prospective field study to evaluate the performance and acceptance of a blood-based HIV self-test in Canada. BMC Public Health. 2021;21:1421. 10.1186/s12889-021-11418-z. pmid:34275450

[17] Ngoc BV, Majam M, Green K, Tran T, Hung MT, Que AL, et al. (2023) Acceptability, feasibility, and accuracy of blood-based HIV self-testing: A cross-sectional study in Ho Chi Minh City, Vietnam. PLOS Glob Public Health. 2023 3(2): e0001438. 10.1371/journal.pgph.0001438

[18] Ndungu K, Gichangi P, Temmerman M (2023) Evaluation of factors associated with HIV self-testing Acceptability and Uptake among the MSM community in Nairobi, Kenya: A cross sectional study. PLoS ONE 18(3): e0280540. 10.1371/journal.pone.0280540

[19] Pant Pai, N., Bhargava, M., Joseph, L., Sharma, J., Pillay, S., Balram, B., & Tellier, P.-P. (2014). Will an Unsupervised Self-Testing Strategy Be Feasible to Operationalize in Canada? Results from a Pilot Study in Students of a Large Canadian University. AIDS Research and Treatment, 2014, 1–8. doi:10.1155/2014/747619

[20] Figueroa C, Johnson C, Ford N, Sands A, Dalal S, Meurant R, et al. Reliability of HIV rapid diagnostic tests for self-testing performed by self-testers compared to health-care workers: a systematic review and meta-analysis. Lancet HIV. 2018; 5(6):e277–e290.

[21] Prazuck, Karon, Gubavu, Andre, Legall, Bouvet, et al. A finnger-stick whole-blood HIV self test as an HIV screening tool adapted to the general public. PloS One. 2016; 11(2):e0146755.

[22] World Health Organization [WHO]. HIV Self-Testing Strategic Framework: A Guide For Planning, Introducing And Scaling Up HIV Testing Services. Geneva 2018.

